# Genetic Associations with Alzheimer’s, Lewy Body and Vascular Neuropathology Reveal Shared and Distinct Biological Pathways

**DOI:** 10.64898/2026.09.16.26362814

**Authors:** Kevin Morgan, Alan Thomas, Keeley J Brookes

## Abstract

**INTRODUCTION:** Alzheimer’s, Lewy body and vascular neuropathology underpin the most common forms of dementia. Large-scale genetic studies tend to rely on clinical diagnoses and therefore may be subject to misdiagnoses and co-neuropathologies that occur.

**METHODS:** Utilising the extension neuropathology data available on the Brains for Dementia Research cohort, this study performed whole-genome association analyses based on common neuropathology present in dementia. Genetic data was generated on the Illumina Neurobooster array and analysed using standard analyses pipelines in PLINK.

**RESULTS:** Several associations were observed in and around the *APOE* locus but only in relation Alzheimer’s neuropathology. Whilst there was a convergence of common gene associations across the different neuropathology groups, there were distinct associations of individual SNPs.

**DISCUSSION:** Different neuropathological outcomes appear to be driven by distinct genetic variants but converge on shared genes and biological processes related to neuronal structure and communication. Understanding these common pathways may provide new insights into mechanisms underlying mixed neuropathology and age-related neurodegeneration.

## Background

Dementia is a heterogenous disorder, with multiple underlying disease pathways leading to the presentation of symptoms. Even within these specific sub-types of dementia, such as Alzheimer’s disease (AD) large heterogeneity is thought to exist, specifically when it comes to genetic aetiology^1^. To address this, instead of using clinical diagnoses to group individuals for genetic association studies an alternative analysis is to explore disease-related neuropathology.

The semi-quantitative measures characterising the underlying neuropathology of dementia may be considered endophenotypic markers bridging the gap between genetic variation and diagnoses. The definition of an endophenotype has recently been updated^2^ “as ‘genetically influenced phenotypes linked to disease or treatment characteristics and their related events.’ To qualify as an endophenotype, the trait must be reliably measured, associated with the disease of interest and mediated by genetic aetiology. The revised framework has broadened genetic association to include causality, pleiotropic effects and non-linear relationships. The definition also encompasses genetically regulated responses to disease-related factors, including environmental risks, illness progression, treatment responses, and resilience phenotypes.^2^ The use of endophenotypes is believed to increase statistical power by reducing phenotypic heterogeneity, using measures closer to gene action and utilising a greater number of samples.

In dementia research, endophenotypes such as levels of phosphorylated Tau (pTau) and Amyloid-beta 42 (Aβ_42_) in cerebrospinal fluid/blood^3,4^, brain structural measurements through MRI^5–8^ and measures of amyloid plaque density and neurofibrillary tangles in the brain have been studied^9,10^.

The Brains for Dementia Research (BDR) cohort is a dataset of longitudinally collected dementia-related assessments, alongside clinical data and post-mortem neuropathology measurements from donated brain tissue for research purposes^11^. Previously DNA has been extracted from BDR brain tissue as used in diagnosis-based gene association studies^12,13^.

In this new analysis on an expanded dataset, genetic analysis of neuropathic features which determine the extent of Alzheimer’s, Lewy Body and vascular neuropathology underlying the dementia diagnosis is carried out. Using established neuropathological scoring systems^14–16^ to assess the level of observed neuropathology contributing to cognitive impairment, a case-control genetic association study compared individuals with high and low neuropathological burden. This approach aimed to strengthen genetic signals by reducing the clinical heterogeneity inherent in dementia diagnoses.

## Methods

### BDR cohort

Cerebellum tissue and blood were sought from BDR over several years to build a DNA bank of the cohort^13^. To date, 1520 DNA samples are contained with the bank, consisting of 1181 deceased and 339 living BDR participants. All blood samples and brain tissue were collected with informed consent as governed by local guidelines at the point of enrolment. Prior to 2019 the study was conducted under the general ethical approval obtained from BDR London – City and East NRES committee 08/H0704/128+5 and completed in accordance with approved guidelines. Since 2019 ethical approval was granted by each brain bank individually under their ethical approval references: MRC London Neurodegenerative Diseases Brain Bank (18/WA/0206), Oxford Brain Bank (23/SC/0241), South West Dementia Brain Bank (18/SW/0029), Newcastle Brain Tissue Resource (08/H0906/136+5) and Manchester Brain Bank (09/H0906/52+5)

### DNA extraction

DNA was extracted from BDR samples using standard phenol-chloroform procedures. Samples were quantified using the Nanodrop 3300 spectrometer to ensure high concentration and quality material was obtained.

### Genotyping

One thousand four hundred and twenty-six BDR DNA samples were genotyped using the Neurobooster Array^17^ (Illumina), capturing 1.825 million genetic variants from the existing Global Diversity Array with an additional 95,273 variants associated with more than 70 neurological conditions or traits. Samples had APOE ε2, ε3 and ε4 isoform determined from the genotyping of SNPs rs7412 and rs429358 using TaqMan assays (Applied Biosystems). This genotype data is available upon request to the BDR coordinating centre or UKBBN.

Raw intensity files (idats) were imported into Illumina’s GenomeStudio_v2.0 for genotype calling. Manual curation of the SNP clustering performed by GenomeStudio algorithms was conducted on all SNPs. SNPs with ambiguous clustering of the three genotypes were removed. Likewise, individual sample signals which lay ambiguously between genotype clusters were also removed. Genotypes from the Neurobooster array were exported from GenomeStudio_v2.0 in PLINK compatible format with chromosomal coordinates aligned to the GRCh37/hg19 assembly.

Quality control on the dataset was conducted using PLINK_v1.9^18^. SNPs with call rates of <95%, a minor allele frequency <5% and genotype frequencies significantly out of Hardy-Weinberg equilibrium (p<1x 10^−6^) were removed from the dataset (n=611,278 variants remaining for analysis). Samples were removed based on biological sex mismatch with reported sex on UKBBN, genotyping rates <95% and deviations from European Caucasian population clustering based on data from 1000genomes^19^.

### Phenotypes

Neuropathology data was obtained for all samples from the UKBBN database, including, Thal staging, Braak Tangle Staging, CERAD; Braak LB staging, Presence of infarct >10mm, and moderate/severe presence of Arteriosclerosis in Occipital White Matter, and Occipital Leptomeningeal CAA.

These neuropathic features were incorporated into scoring systems which underlie the likelihood the level of neuropathology observed is responsible for the cognitive symptoms presented. ABC scoring was used to determine level of Alzheimer’s neuropathology^14^; whilst VCING scoring was used to determine levels of dementia-relevant vascular neuropathology^15^. Braak staging^16^ was utilised as the measure of Lewy body neuropathology burden (Supplemental Material: Table S1).

Categories of absent, low, moderate and high were generated according to the ABC^14^ and VCING^15^ systems. Lewy body Braak staging was stratified into absent (Stage 0), low (Stages 1–2), moderate (Stages 3–4), and high (Stages 5–6) tiers to capture the structural progression of neuropathology, mapping directly from isolated lower brainstem pathology (low) through transitional limbic involvement (moderate) to widespread neocortical burden (high).

Binary phenotypes for genetic analysis were classified with individuals with absent or low neuropathology as controls, whilst those scoring moderate or high levels of neuropathology were regarded as cases. If measures were not recorded/available from UKBBN database, neuropathology was denoted as missing.

### Analysis

Logistic regression in PLINK_v1.9^18^ was conducted for all three neuropathologies independently taking into consideration standard dementia covariates: age at death; biological sex and number of APOE ε4 isoforms.

Principle component, Manhattan/Q–Q and upset plots were generated using R packages: “eulerr”^20^, “qqman”^21^ and “UpSetR”^22^ respectively.

G:profiler SNPense^23^ was used to identify associated genes with genetic variants, and STRING^24^ was used to assess gene lists for shared biological functions and networks.

## Results

### Genotyping

A total 1,901,182 variants were clustered in GenomeStudio_v2.0. After visual inspection those with poor clustering were removed leaving 1,816,257 genetic variants for export. Seven samples were removed at this stage due to yielding very poor genotyping results.

Using PLINK_v1.9^18^, samples were checked for biological sex inferred by genotyping, 13 samples were removed from the dataset as inferred sex did not match stated biological sex in UKBBN database. Twenty-six samples were duplicated, either across batches of genotyping or from DNA taken from brain and blood. Relatedness analysis in PLINK_v1.9 indicated that duplicates were >0.999 identical in genotype calls. Duplicates with the lower genotyping rate were removed. A further eight samples were removed to due high relatedness (>0.87) between different sample IDs. Two samples were removed due to self-reported ethnicity other than Caucasian; a further sample was removed after inspection of PCA plot alongside 1000G population data (Supplementary Material: Table S2).

### BDR Sample

Of the cohort with genetic data available 1057 participants had sufficient neuropathology data to determine presence of dementia neuropathology (Figure 1). Whilst 29.4% of the samples were determined to have no or low dementia-related pathology; 44.7% of the cohort presented with a single dementia-related neuropathology, and 25.9% of the participants were determined to have mixed or multiple neuropathologies.

**Figure 1:**
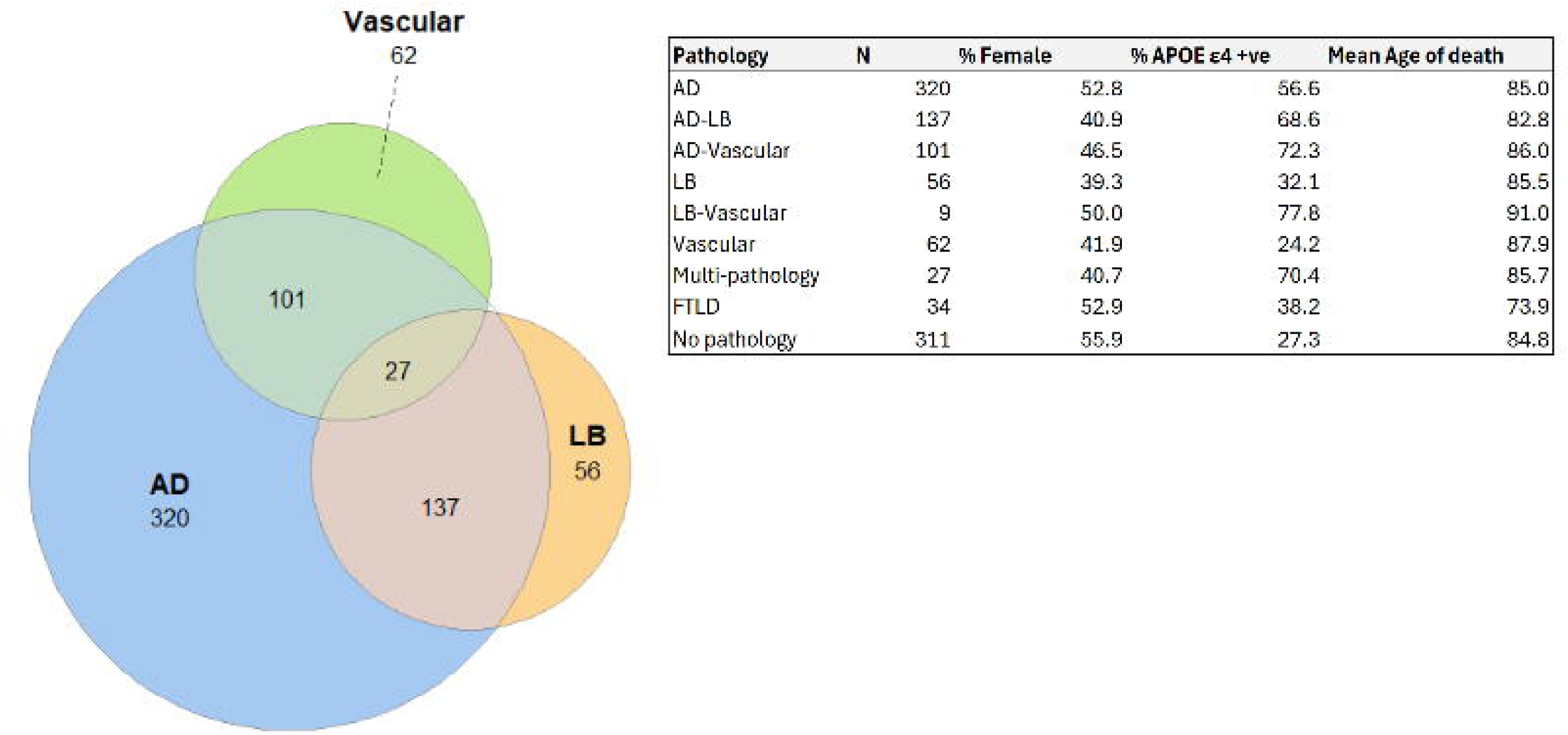
Venn diagram of the three neuropathologies investigated in the study, Demonstrating the significant overlap of these neuropathologies present in BDR participants. The accompanying table details the sample size and key co-variants in dementia for each neuropathology group.

Clinical diagnoses did not always match up with the neuropathology data observed; for example, the most frequent clinical diagnosis was for AD and was assigned to 324 BDR participants; only 125 were confirmed by the presence of significant (moderate/high) Alzheimer’s neuropathology only, along with 118 participants which had Alzheimer’s pathology alongside other dementia-related neuropathologies (Lewy body and/or vascular).

Only 193 of the 345 (55.9%) participants described as cognitively normal at the last interview prior to death were absent of any dementia-related neuropathology. No neuropathology data were available for 3.8% (n=13), whilst the remaining 39.4% had significant dementia-related neuropathology. The most common neuropathology observed was Alzheimer’s, with 62 participants displaying amyloid and tau pathology exclusively, and a further 21 participants presenting with Alzheimer’s pathology alongside Lewy body and/or vascular neuropathology. Comparisons with the cognitively normal control group yielded no significant differences between the proportion of biological females present in each group nor the presence of APOE ε4 or ε2 isoforms. The only significant difference between the cognitively normal groups was the age-at-death, where there was an average age of 85.4 years (±8.4) for the no neuropathology sub-group and an average age of 90.4 (±7.3) for those with observed neuropathology. Analysis of the time-period from time of last interview schedule to death, was not significantly different between the groups (p=0.49) with an average time of 12.6 (±11.9) and 13.5 (±13.2) months for the no neuropathology and neuropathology present groups respectively.

Conversely there were 86 participants who received a dementia diagnosis, where no significant dementia associated pathology was observed. Average age-at-death was calculated at 85.1 (±10.9) years, 50% of the group were female, with proportions of APOE ε4 or ε2 isoforms at 37.2% and 17.8% respectively.

### Association results

Logistic regression analysis of binomial presence/absence of pathology identified ∼30,000 variants to be associated with each pathology at a nominal significance level (p<0.05, Figure 2). More stringent cut-offs were applied but given the sample size no findings withstood the multiple correction to the study-wide and genome-wide level, except for a single SNP in the *APOE* gene associated with Alzheimer’s neuropathology. The predominant neuropathology observed was Alzheimer’s accounting for 50% of dementia-related neuropathologies in this cohort. Ratios of controls to cases was 1.4:1 for Alzheimer’s neuropathology; whereas with both Lewy body and vascular neuropathology the ratios were 3.3 and 3.6:1 respectively.

**Figure 2:**
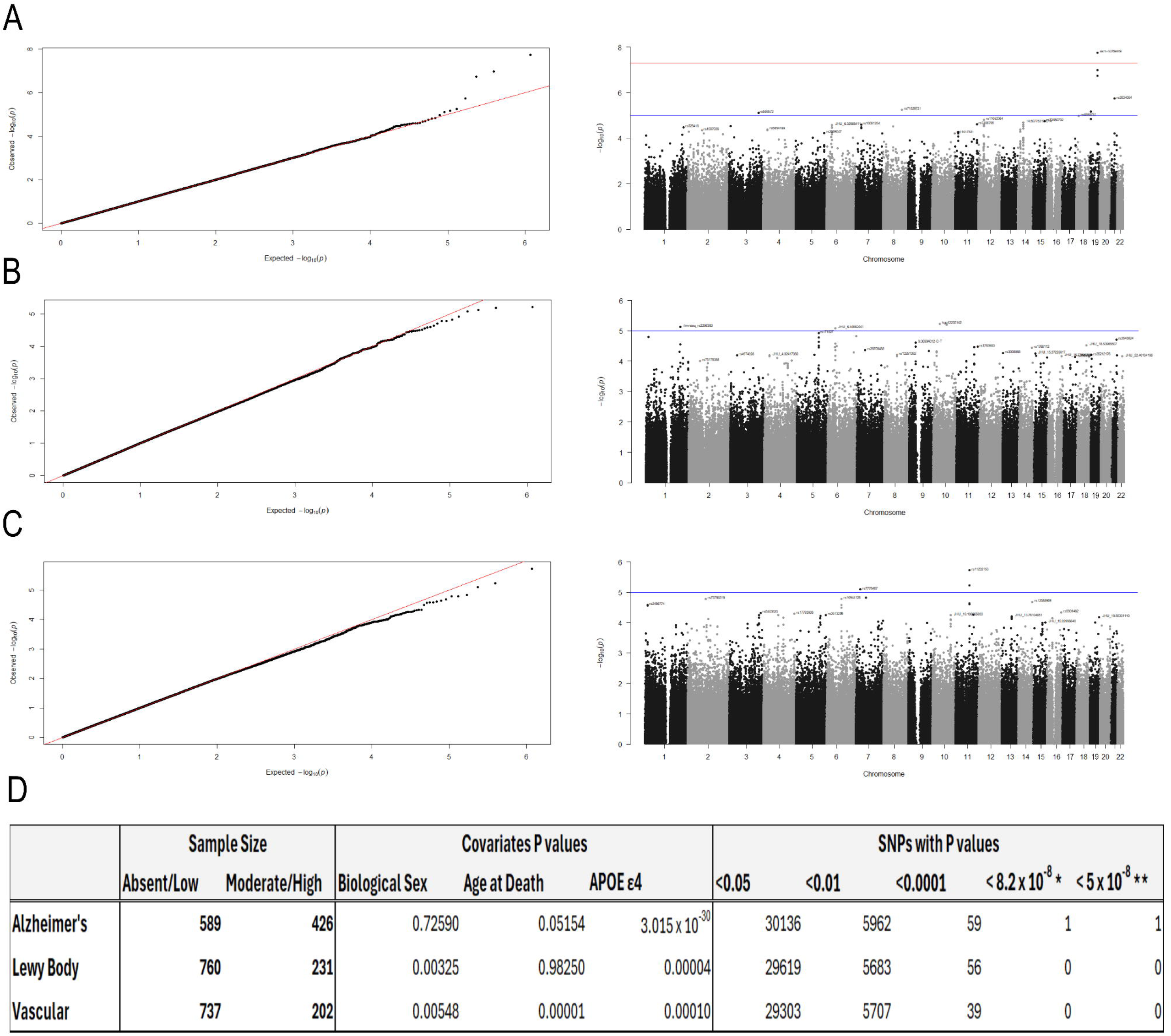
QQ and Manhattan plots of association results (including co-variates) for each neuropathology A: Alzheimer’s; B: Lewy body; C: Vascular. D is a summary table of the sample seizes for each analysis, association of co-variates and variants captured at each significance level.

Covariates of biological sex, age-at-death and number of APOE ε4 isoforms were included in all analyses and highlighted several differences amongst the neuropathology groups. The number of APOE ε4 isoforms were highly associated with Alzheimer’s neuropathology with each additional ε4 isoform increasing risk of having significant neuropathology by 4.6 times (p=3.02 x 10^−30^). However, this risk was greatly diminished for both Lewy body and vascular neuropathology with additional ε4 isoforms increasing risk by 1.6 times (p=3.94x10^−5^; p=0.0001 respectively). Age-at-death was only found to be associated with vascular neuropathology (p=1.3x10^−5^), and biological sex indicated that females had 40% decreased risk compared to males of having high levels of Lewy body and vascular neuropathology (p=0.003 and p=0.005 respectively).

### Alzheimer’s neuropathology

The top three significant SNPs for Alzheimer’s neuropathology were all on chromosome 19 and within ∼13kb upstream of the rs429358 *APOE* isoform SNP. The top hit was the only one to remain significant after study/genome-wide correction; rs769449 is an intronic variant within the *APOE* gene, 1939bp upstream to the rs429358 *APOE* isoform SNP. Additional top hits (p<0.0001) were genes involved with neuronal survival, and innate immunity/inflammation with a number associated with cancer (Supplementary Material: Table S3). For example, *MUC16* (6.69x10^−6^) associated via SNP rs8111512, and several associated variants (rs2297995, rs11157734, rs2355885) in the *L2HGDH* gene (p<2.61x10^−5^) and *ADCYAP1R1* gene (rs10081254, rs113015906; p<3.325x10^−5^).

Out of the 76 genes listed as candidates for AD compiled by ADSP Gene Verification Committee, 22 were also identified with nominal significance in this study (Supplemental Material: Table S6). In addition, previously associated genes such as *DLG2, MEF2C, CD33, RIN3*, were also identified as associated with AD-related pathology (p<0.05).

STRING^24^ analyses of the 7433 genes identified from the variants with nominal significance (p<0.05), indicated a highly significant over representation of genes involved with neuronal development and neurogenesis (p<1.4x10^−11^) biological processes. The top molecular function identified was calcium and metal ion transmembrane transporter activity (p<7.69x10^−6^). With immunoglobulin and fibronectins identified as the top protein domains overrepresented in this dataset (p<3.64x10^−6^).

### Lewy body neuropathology

Whilst no variants surpassed the genome/study-wide multiple testing correction, the top association for presence of Lewy body neuropathology was kgp12255142 a variant on chromosome 10 within the *PLD5P1* gene (p=5.97x10^−7^). Other genes with highly significant variants within them include *KCNMA1* (rs35366821; p=6.4x10-6), *CACNA1S* (rs2296383, rs10159219; p<2.78x1-05), *FBN2* (rs171527, rs331069, rs469722; p<3.11x10-5), KAZN (rs7535502; p1.61x10-5) and *VEGFA* (rs3024994; p=2.62x10-5).

Other known Lewy body disease associated genes such as *SNCA* (rs1442144, p=0.038) and *USP13* (rs6443665, rs16830685 & rs62291845, p <0.05) were also detected in this study (Supplementary Material: Table S4).

Given the high prevalence of mixed neuropathologies not often detected in clinically diagnosed studies, a comparison of associated findings with the ADSP gene list was conducted and 23 variants within these genes were nominally significant with Lewy body neuropathology (Supplemental Material: Table S6).

STRING^24^ analyses of the 7158 genes identified from the variants with nominal significance (p<0.05), identified an over representation of genes involved in cell morphogenesis (p<4.73x10^−10^), and with molecular functions of gated and ion channel activity (p<5.59x10^−8^). As with the Alzheimer’s neuropathology genes, there was a significant over representation of genes immunoglobulin (p=6.06x10^−6^) and fibronectin (p=4.62x10-5) protein domains.

### Vascular neuropathology

Again, no variants surpassed the genome/study-wide correction for association with vascular neuropathology. The top hit was an intergenic SNP rs11232153 on Chr11 (p=1.89 x 10^−6^). Other top variants located within known genes included rs10944128 (*NT5E*; p=1.63 x 10-5), rs6909797 (*SNX14*; p=2.63x10-5), rs12212560 (*SYNCRIP*; p=3.22x10-5) and *KAZN* was implicated with two SNPs rs2486774 and rs2486762; p<2.71x10-5).

Genes previously associated with Vascular dementia such as *ASTN2* was identified in this study with SNP rs10983609 (p=0.0088), however *MTHFR* was not found to be nominally significant (Supplementary Material: Table S5).

Inspection of variants against the ADSP gene list observed 22 genes of the Alzheimer’s disease genes were associated via Vascular neuropathology associated variants at the nominal significance level (Supplemental Material: Table S6).

Seven thousand, one hundred and thirty-eight genes were identified from the vascular neuropathology analysis and interrogated with STRING^24^ for biological pathway over representation. Comparable to the gene list identified from the Lewy body neuropathology, cell morphogenesis was the top biological process represented by the genes identified (p<1.99x10^−9^) with ion channel activity as the top molecular function (p<1.61x10^−7^). As observed in our previous neuropathology analyses, the immunoglobulin domain was again the most significantly overrepresented protein domain among the associated genes (p< 2.08 × 10^−^□).

### Comparison of data sets

Both associated genetic variant and gene lists were compared between the three neuropathology analyses (Figure 3). Pairwise results suggested that around 1400–1600 variants were common between neuropathologies (4–5% of the nominally significant variants observed). There was a greater commonality between gene lists with around 3000 genes identified between neuropathology lists. Shared SNPs between all three neuropathology groups totalled 99, whilst the number of genes in common totalled 1947.

**Figure 3:**
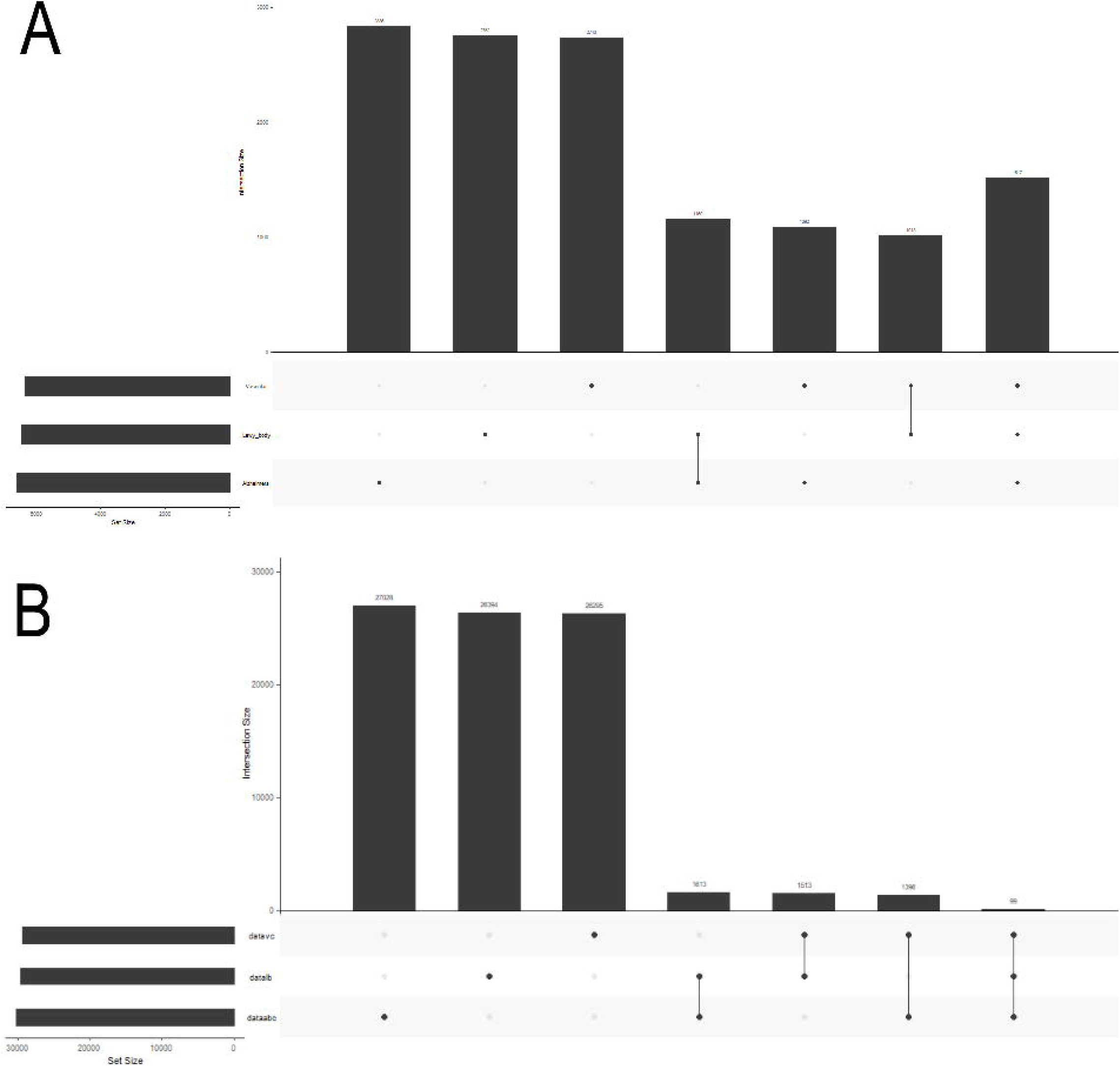
Upset plots indicating the number of unique and common A: Genes and B: variants identified at the nominal significance level (p<0.05) across the neuropathology groups. Demonstrating shared genes within different pathologies though distinct genetic variation.

A prominent gene identified was *GABRG3* implied by three SNPs within this locus being associated at the nominal level across all three neuropathologies investigated. Other SNPs of interest that were associated in all three dementia-related neuropathologies included SNP rs35561044 in the chr 21 gene *DSCAM*; rs137190 in *SEZ6L* a substrate of BACE1; rs11188301 in *SORBS1*, and rs1230106 in *ARHGAP27*, in close linkage disequilibrium with the *MAPT* gene. In all these 99 variants tagged 27 protein coding genes.

The gene list of 1947 genes that were commonly implicated by nominally associated SNPs across the neuropathologies, contained 939 protein coding genes, with 87% (n=817) forming a single cluster in STRING pathway analysis with an enrichment p value of 1x10^−16^. (Figure 4). As with the single neuropathology analysis top biological functions include cell morphogenesis and neuronal development, and a significant over representation of immunoglobulin and fibronectin protein domains.

**Figure 4:**
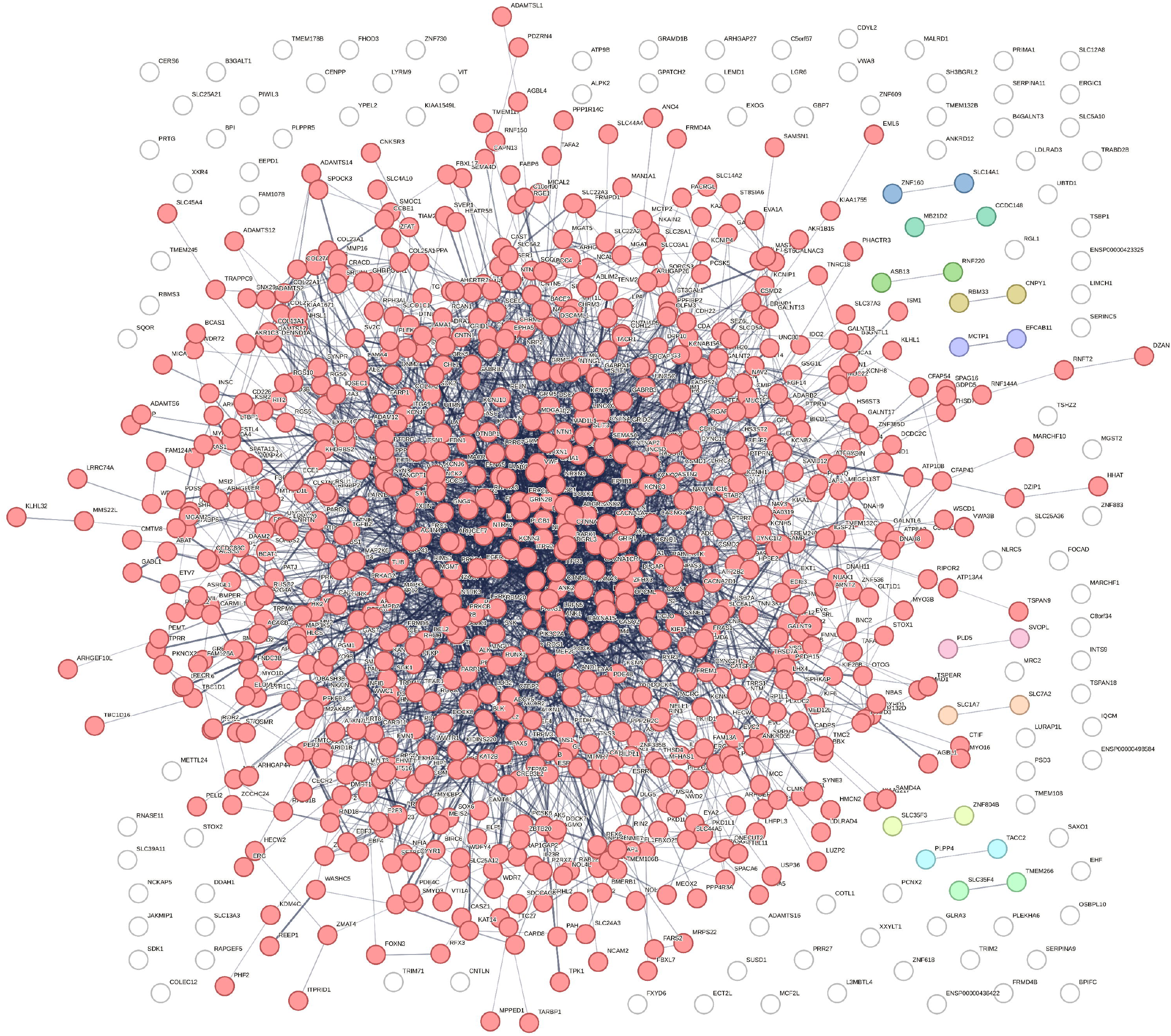
K-means clustering performed in STRING of genes commonly identified in each neuropathology, indicating that there may be a shared underlying biological pathway underlying brain vulnerability to dementia.

Eight of the genes commonly identified across the neuropathology groups were amongst the ADSP gene list (Supplemental Material S6), alongside other known associated genes from previous GWAS including *MEF2C, RIN3* and *DLG2*. Interesting in all these cases, the genes were implicated by different SNPs associated with each neuropathology, and in some cases (*EPHA, RIN3, DLG2 & ANK3*) had different directions of effect size. None of the SNPs identified with Alzheimer’s neuropathology were found at the nominal significance level in either Lewy body or vascular neuropathology and vice versa, though there was some cross-over between Lewy body and vascular neuropathology with top associated SNPs in *ICA1* (rs6463769), *ABCA1* (rs2487052) and *RIN3* (rs10142182), also found in the corresponding neuropathology though at different significance levels and in the case of *ICA1* with a different direction of effect.

## Discussion

This study sought to explore the genetic associations with differing dementia neuropathologies. The BDR cohort undergoes extensively neuropathology assessment at post-mortem identifying the underlying neuropathology to account for dementia symptoms.

Exploration of BDR participant clinical diagnoses and neuropathology present emphasises previously documented misdiagnosis^25^, with the presence of Alzheimer’s disease neuropathology present in only a third of cases identified as Alzheimer’s disease in life. Likewise, almost half of cognitively normal at-death participants were found to have significant dementia-related neuropathologies^26^.

It was hypothesised that these participants might have died at a younger age prior to their symptom onset and therefore would have a lower age-at-death than their pathology absent counterparts. However, whilst the average age-at-death was 85.8 years for cognitively normal participants without pathology, the group where dementia-relevant pathologies were observed had an average age of 90.4 years. This would suggest that instead of dying before symptom onset, this group may demonstrate resilience against cognitive decline despite the presence of dementia-related neuropathology, and merit further study.

The most associated SNP with Alzheimer’s neuropathology, and the only variant to remain significant at the genome-wide level in the investigation was rs769449, and intronic SNP in *APOE*. This SNP has been found to be highly significant in other genome-wide association studies (GWAS) for AD diagnosis^27,28^. The presence of the A-allele is associated with the presence of tau pathology, via associations with levels of pTau in both CSF and blood plasma^3,4,29^. This aligns with the current study as for moderate/high classification in the ABC scoring system, Tau Tangle Braak staging III must be present alongside amyloid neuropathic features (Supplemental Material S1). It is unknown if this is the “causal” variant given its high linkage disequilibrium (r^2^>0.8) with the coding SNP rs429358, which determines the presence of APOE ε4 isoform^30^, however according to GTEx^31^ database, rs769449 is an expression trait loci for *TOMM40* and therefore may have influence on biological outcomes.

Furthermore, our genetic association analysis identified a novel triad of loci—*MUC16, L2HGDH*, and *ADCYAP1R1* with Alzheimer’s neuropathology. *ADCYAP1R1* encodes the primary receptor for the neuroprotective peptide PACAP, which is a promising new modifying drug for AD^32^. PACAP increases α-secretase activity promoting the non-amyloidogenic pathway^33^. Both SNPs identified in this study are intronic within the *ADCYAP1R1* locus and therefore might affect transcriptional regulation of the gene and consequently have knock-on effects on the seemingly beneficial PACAP pathway.

Significantly associated genes with Braak staging for Lewy body neuropathology represented ion channel activity molecular functions. Disruption of ion-channel-mediated calcium signalling is increasingly recognized as a contributing mechanism in neurodegeneration. The identification of *KCNMA1* and *CACNA1S* therefore points towards pathways involved in cellular excitability and calcium regulation, processes that are critical for synaptic function and neuronal survival. Whilst *CACNA1S* is mainly expressed in skeletal muscle rather than the central nervous system, low muscle mass is becoming recognised as a risk factor for dementia^34,35^ and the gene itself has been associated with episodic memory^36^. *KCNMA1* encodes the pore-forming alpha subunit of the BK (Big Potassium) channel, has been observed in previous GWAS, specifically with age-of-onset^37^.

Variants associated with the VCING measure of vascular neuropathology included those in genes *NT5E, KAZN, SYNCRIP*, and *SNX14*. Crucially, *NT5E* (encoding ecto-5’-nucleotidase/CD73) acts as a primary vascular driver within this network; mutations within this gene are known to increase arterial calcifications^38^, which in turn has been shown to be a risk factor for cognitive decline^39,40^. This calcification restricts blood flow in the brain and subsequently creates a hypoxic environment and small vessel disease. A recent GWAS for small vessel disease demonstrated a number over genetic overlaps with the findings presented here for vascular neuropathology^41^.

Despite differences in the specific neuropathological phenotypes examined and genetic associations, STRING enrichment analysis consistently identified pathways related to neuronal structure, cell morphogenesis, ion-channel-mediated signalling and adhesion molecules containing immunoglobulin and fibronectin domains. Roughly 25% of genes identified in this study were common across the three neuropathology groups. Unsurprisingly these genes clustered into a single protein network potentially representing a common vulnerability behind dementia subtypes. Consequently, this network may highlight potential drug targets for therapeutic development. Many of the large AD GWAS studies, including those feeding into the ADSP AD gene list, define cases clinically. However, as observed here, co-neuropathology in clinically diagnosed groups is likely. Out of the 712 participants with notably neuropathology investigated here, 38% had co-neuropathology. The overlap between genes identified in the present neuropathology-specific analyses and those previously implicated by ADSP may reflect the substantial burden of mixed neuropathology present within large Alzheimer’s disease GWAS cohorts, whereby associations attributed to AD risk may partly capture genetic influences on co-occurring Lewy body and vascular neuropathology.

However, an intriguing observation is the SNP heterogeneity between commonly associated genes across the neuropathology groups and that some loci have opposite directions of effect. This suggests pleiotropic gene effects and potentially distinct biological mechanisms underlying individual neuropathological processes, and an important avenue to explore.

A recognised limitation of this study is the sample size, and the extent of co-neuropathologies. Despite the relatively small sample size in comparison to typical genome-wide association cohorts, the neuropathic detail reduces the likely heterogeneity of these much larger datasets^42^. However, owing to sample size constraints, analyses were conducted according to the presence or absence of individual pathologies rather than examining all pathological combinations separately. Consequently, while analyses were stratified by the presence or absence of individual neuropathologies, residual confounding from co-pathology remains possible.

An example of this could be the association with the *APOE* gene. Whilst the presence of the risk isoform of APOE was considered, several associations were noted in and around the *APOE* gene locus with Alzheimer’s neuropathology (3 variants in/near *APOE*, 8 in *NECTIN2* and 2 in *TOMM40)*, whilst variants this region were not identified as significantly associated with either Lewy body or vascular neuropathology (p<0.05). However, covariate analysis indicated that the number of APOE ε4 isoforms were still associated with these neuropathologies, though to a lesser extent, which could be attributed to the number of participants that also have Alzheimer’s neuropathology.

To maximize statistical power and overcome the sample size constraints in the four-tier ordinal classification systems present of pathology was dichotomized into a binary case-control framework. While endophenotypic analyses ideally leverage multi-categorical or continuous quantitative metrics this would have under-powered the regression models. Transforming the metric into a dichotomous trait consolidates statistical power within a rigorous logistic or case-control regression framework, mitigating the risk of type II errors while still reliably capturing the fundamental genetic liability associated with the neuropathology.

This study did not use the standardized McKeith scoring system for Lewy body disease categorization^43^, which is traditionally cross-referenced alongside ABC and VCING criteria to establish the clinicopathological probability of dementia. To maximize sample size and preserve statistical power, the Lewy Body Braak staging data was used, which was more comprehensively available across BDR. While this prevents a direct mapping onto the specific McKeith likelihood matrices, the strict hierarchical overlap between subcortical/cortical Braak stages (1–6) and McKeith tiers (Brainstem, Limbic, Diffuse Neocortical) mitigates this limitation. Consequently, the findings remain highly representative of regional Lewy pathology progression.

Furthermore, our choice of Lewy Body Braak staging over the McKeith system aligns with the primary objective of this study, which focuses strictly on mapping the structural presence and anatomical progression of neuropathy. The McKeith consensus matrix is fundamentally designed as a clinicopathological tool; it explicitly incorporates Alzheimer’s Tau Braak staging to weigh competing pathologies and determine which is most likely responsible for a patient’s cognitive symptoms. Because our analysis aims to identify genetic variation associated with structural burden of Lewy body neuropathology alongside independent markers like ABC and VCING scoring, using Lewy body Braak staging was more appropriate.

In conclusion, this study identified both shared and distinct genetic associations across Alzheimer’s, Lewy body and vascular neuropathological phenotypes. Despite limited overlap at the SNP level, the convergence of associated genes and enriched pathways across neuropathological phenotypes suggests that Alzheimer’s disease, Lewy body disease and vascular neuropathology may share aspects of a common biological framework centred on neuronal structure, connectivity and excitability. The recurrent identification of genes involved in ion transport, cell morphogenesis and adhesion-related protein domains supports a model in which disruption of neuronal communication and network integrity contributes to multiple forms of age-related neuropathology. Further studies integrating genetics with molecular and neuropathological data will be required to determine whether these shared pathways represent common mechanisms of neurodegeneration or distinct processes that ultimately converge on similar clinical outcomes.

## Data Availability

All data produced in the present study are available upon request to the BDR coordinating centre

## Acknowledgements

We would like to gratefully acknowledge all donors and their families for the tissue provided for this study. Human post-mortem tissue was obtained from the Southwest Dementia Brain Bank, London Neurodegenerative Diseases Brain Bank, Manchester Brain Bank, Newcastle Brain Tissue Resource and Oxford Brain Bank, members of the Brains for Dementia Research (BDR) Network.

## Consent Statement

All blood samples and brain tissue were collected with informed consent as governed by local guidelines at the point of enrolment.

## Conflicts

The authors declare no conflict of interests and have no disclosures.

## Funding Sources

This work was funded by an Alzheimer’s Society Major Project award to KJB, KM and AT, entitled “Adding value to the Brains for Dementia Research cohort with additional genetic data” (AS-PG-19b-001).

## Acknowledgements

We would like to gratefully acknowledge all donors and their families for the tissue provided for this study. Human post-mortem tissue was obtained from the Southwest Dementia Brain Bank, London Neurodegenerative Diseases Brain Bank, Manchester Brain Bank, Newcastle Brain Tissue Resource and Oxford Brain Bank, members of the Brains for Dementia Research (BDR) Network. We would also like to acknowledge UCL Genomics where the Neurobooster Genotyping was conducted.

## Funding

Funding awarded to Dr Keeley Brookes as PI, and co-PI Prof Kevin Morgan and Prof Alan Thomas by Alzheimer’s Society Major Project Award entitled: “Adding value to the Brains for Dementia Research cohort with additional genetic data” (AS-PG-19b-001).

## Competing Interest

The authors report no other competing interests.

## Author Contributions

All authors contributed to the study conception and design. All authors read and approved the final manuscript. Keeley Brookes conducted laboratory work, analysis of the data and manuscript writing.

## Notes

### Competing Interest Statement

The authors have declared no competing interest.

### Author Declarations

Prior to 2019 the study was conducted under the general ethical approval obtained from BDR London City and East NRES committee 08/H0704/128+5 and completed in accordance with approved guidelines. Since 2019 ethical approval was granted by each brain bank individually under their ethical approval references: MRC London Neurodegenerative Diseases Brain Bank (18/WA/0206), Oxford Brain Bank (23/SC/0241), South West Dementia Brain Bank (18/SW/0029), Newcastle Brain Tissue Resource (08/H0906/136+5) and Manchester Brain Bank (09/H0906/52+5)

